# Eligibility to COAPT trial in the daily practice: a real-world experience

**DOI:** 10.1101/2024.02.27.24303471

**Authors:** Edoardo Zancanaro, Nicola Buzzatti, Paolo Denti, Nicolò Azzola Guicciardi, Enrico Melillo, Fabrizio Monaco, Eustachio Agricola, Francesco Ancona, Ottavio Alfieri, Michele De Bonis, Francesco Maisano

**Author notes:** **Address for correspondence:** Edoardo Zancanaro MD, Department of Cardiac Surgery, San Raffaele Scientific Institute, Milan, Italy.; mail. No competing interests and no contributorship.

## Abstract

**Background:** The COAPT Trial was the first ever to demonstrate a survival benefit in treating functional mitral regurgitation (FMR). That was achieved through transcatheter mitral repair in selected patient. The exact proportion of patients fulfilling COAPT selection criteria in the real-world is unknown.

**Objectives:** to assess the applicability of COAPT criteria in real-world and its impact on patients’ survival.

**Methods:** We assessed the clinical data and follow-up results of all consecutive patients admitted for FMR at our Department between January 2016 and May 2021 according to COAPT eligibility. COAPT eligibility was retrospectively assessed by a cardiac surgeon and a cardiologist.

**Results:** Among 394 patients, 56 (14%) were COAPT eligible. The most frequent reasons for exclusion were MR<=2 (22%), LVEF <20% or >50% (19%), and non-optimized GDMT (21.3%). Among NON-COAPT patients, weighted 4-year survival was higher in patients who received MitraClip compared to those who were left in optimized medical therapy (91.5% (CI: [0.864, 0.96] vs 71.8 % (CI = [0.509, 0.926]) respectively, p=0.027)

**Conclusions:** Only a minority (14%) of real-world patients with FMR referred to a tertiary hospital fulfilled the COAPT selection criteria. Among NON-COAPT patients, weighted 4-year survival was higher in patients who received MitraClip compared to those who were left in optimized medical therapy (91.5% (CI: [0.864, 0.96] vs 71.8 % (CI = [0.509, 0.926]) respectively, p=0.027)

**Condensed Abstract:** In the present real-life single center experience, only a small proportion of patients with functional mitral regurgitation were COAPT-like. Non-COAPT like patients treated with MitraClip experienced improved survival compared to those left in medical therapy and similar survival compared to patients treated with MitraClip fulfilling COAPT criteria. While these findings require further validation, the numerous patients currently referred to percutaneous repair outside the COAPT criteria should not be denied intervention but should receive a tailored Heart-Team evaluation. Further refinement of patients selection for transcatheter mitral valve repair and longer follow-up remain necessary.

**Key messages:** **What is already known about this subject?** It is established the efficacy of TEER in case of FMR and the results form the highly selective COAPT trial.

**What does this study add?** This is the first article that prove the applicability of COAPT in real practice and shows the good outcomes also in patients excluded by the trial.

**How might this impact on clinical practice?** This may help the operator not to exclude patients with anatomy and clinical features considered not fit from one of the most important trials in the last 10 years on TEER and medical therapy.

## Introduction

Functional mitral regurgitation (FMR) is frequently present in heart failure (HF) patients with idiopathic or ischemic dilated cardiomyopathy(1), and it is associated with a dismal cardiovascular prognosis(2–4).

Percutaneous mitral valve repair with MitraClip is a consolidated therapeutic strategy for HF patients with clinically relevant FMR. The recent Cardiovascular Outcome Assessment of the MitraClip Percutaneous Therapy for Heart Failure Patients with Functional Mitral Regurgitation (COAPT) was the first-ever randomized trial to demonstrate a survival benefit in treating secondary MR. Such results were achieved by treating patients with MitraClip plus guidelines-directed medical therapy (GDMT) compared with GDMT only(5). Notably, a concomitant and similar trial, the Multicentre Study of Percutaneous Mitral Valve Repair MitraClip Device in Patients with Severe Secondary Mitral Regurgitation (MITRA-FR), provided completely opposed results, ^6^ raising several uncertainties on the matter.

The conundrum to put together the conflicting results of the COAPT and MITRA-FR is still to be fully understood. A strict patient selection in the COAPT trial has been the prevalent explanation for the different outcomes between the two studies. Indeed, in the COAPT the majority of screened patients were refused (911/1576, 58%) with enrollment requiring 5 years to complete across 89 centers. By comparison, the MITRA-FR screening failure rate was 32%, with enrollment completed in 3.5 years at 34 centers.

Today, two main questions still remain unanswered: how many and which patients can achieve COAPT-like outcomes from the MitraClip procedure? How should we best manage patients not falling within the COAPT criteria? To try to improve our knowledge on the matter, we sought to conduct an extensive retrospective analysis across the whole landscape of patients affected by FMR admitted at our Department, assessing their eligibility for COAPT as well as their 4-year survival according to COAPT eligibility and to the actual treatment received.

## Methods

### Study population and data collection

All patients admitted to the Department of Cardiac Surgery at San Raffaele Scientific Institute between January 2016 and May 2021 with the diagnosis of FMR ≥ 3+ were retrospectively selected for the present study. At the time of admission, all patients underwent Heart-Team discussion and prospective in-hospital data collection into a dedicated database for research purposes.

The medical file of each patient was retrieved and assessed for eligibility for the COAPT trial based on the published inclusion/exclusion criteria by the consensus of one hybrid cardiac surgeon (MitraClip operator) and one echocardiographist. Echocardiographic images were also retrieved and assessed when needed. Follow-up data were retrieved by the Institution’s out-patient clinic or performed by telephone calls.

Ethics approval for the study was obtained from our institution’s ethics committee.

### Study endpoints

The primary endpoint of the study was to determine the number of patients who were eligibible for the COAPT trial and to describe the reasons for ineligibility.

The secondary endpoint was to compare 4-year survival of patients according the treatment received.

### Echocardiographic assessment

All patients underwent baseline transthoracic and transesophageal echocardiograms. Transthoracic echocardiogram was then performed after percutaneous repair with MitraClip before discharge and during follow-up visits. MR was graded following current European recommendations(6). A multiparametric approach based on the evaluation of quantitative parameters [effective regurgitant orifice area (EROA) and regurgitant volume (R Vol)] and also semi-quantitative parameters [Vena contracta width (VC) and Color-jet Area], was used to assess MR severity. Right ventricle systolic function was evaluated with tricuspid annular plane systolic excursion (TAPSE), measured with M-mode modality from the tricuspid annular longitudinal excursions in apical 4-chamber view, and with systolic (s’) wave of tissue Doppler velocity imaging (TDI), of basal RV free wall from 4-chamber view. Moderate-to-severe RV systolic dysfunction was defined as TDI s’ wave < 8 cm/s and TAPSE<13mm.

### Statistical analysis

Normally distributed continuous variables were expressed as mean±standard deviation (SD), and compared using the t-test. Continuous variables not normally distributed were expressed as median and interquartile range [IQR] and compared using the Wilcoxon signed-rank test. Categorical variables were reported as absolute number and percentages and compared using the χ2 test or Fisher’s exact test, as appropriate.

To adjust for imbalance in baseline characteristics between different groups, the IPWT technique, based on propensity score matching method, was employed ^8^. Propensity score was estimated running multiple logistic models including various combinations of pre-operative variables. The best balance was achieved with the following 10 variables: age at operation, sex, chronic kidney disease, glomerular filtration rate (GFR), atrial fibrillation, tricuspid regurgitation (TR), left ventricle ejection fraction (LVEF), left ventricle end-diastolic volume (LVEDV), left ventricle end-systolic volume (LVESV) and RV s’-TDI.

Then weights were calculated based on the propensity score method and were trimmed at 95th percentile to ameliorate efficiency and reduce the influence of outlying observations. Balance of confounders between groups was assessed calculating absolute standardized mean differences (ASMD) for the original and weighted sample. Balance was considered good for ASMD <0.10 and sufficient for ASMD <0.20.

Kaplan-Meier weighted survival curves related to all-cause death at 1-year of follow-up were computed and compared using the log-rank test (Cox-Mantel test). A Cox proportional weighted hazard model was employed to assess predictors of mortality. All covariates with p-value < 0.1 at univariate analysis were inserted in a stepwise model. To assess proportional hazard assumption was employed Schoenfeld residuals test. Cumulative Incidence Function (CIF) was computed for cardiac death with non-cardiac related as competing risk. Fine & Gray model was employed to compare treatment. All hospitalizations for heart failure within the first year of follow-up were evaluated and the incidence rate for each group as well as the incidence rate ratio (IRR) between groups were calculated. A joint frailty model was performed to account for correlated events and competing risk of death. Generalized estimation equation analysis was employed to assess change over time of medical therapy to model the possible dependency among repeated measurements. These methods were applied because of the possible intra-patient correlations.

A two-sided p value of less than 0.05 was considered statistically significant. Statistical analysis was performed using Stata Software (Statacorp, LLC, TX, USA; Version 15).

## Results

Out of 394 consecutive patients screened with diagnosis of FMR ≥ 3+, 338 (86%) were found ineligible and 37 (14%) eligible for COAPT trial. Central illustration depicts the patients flowchart based on observed major COAPT inclusion/exclusion criteria.

**Central illustration.**
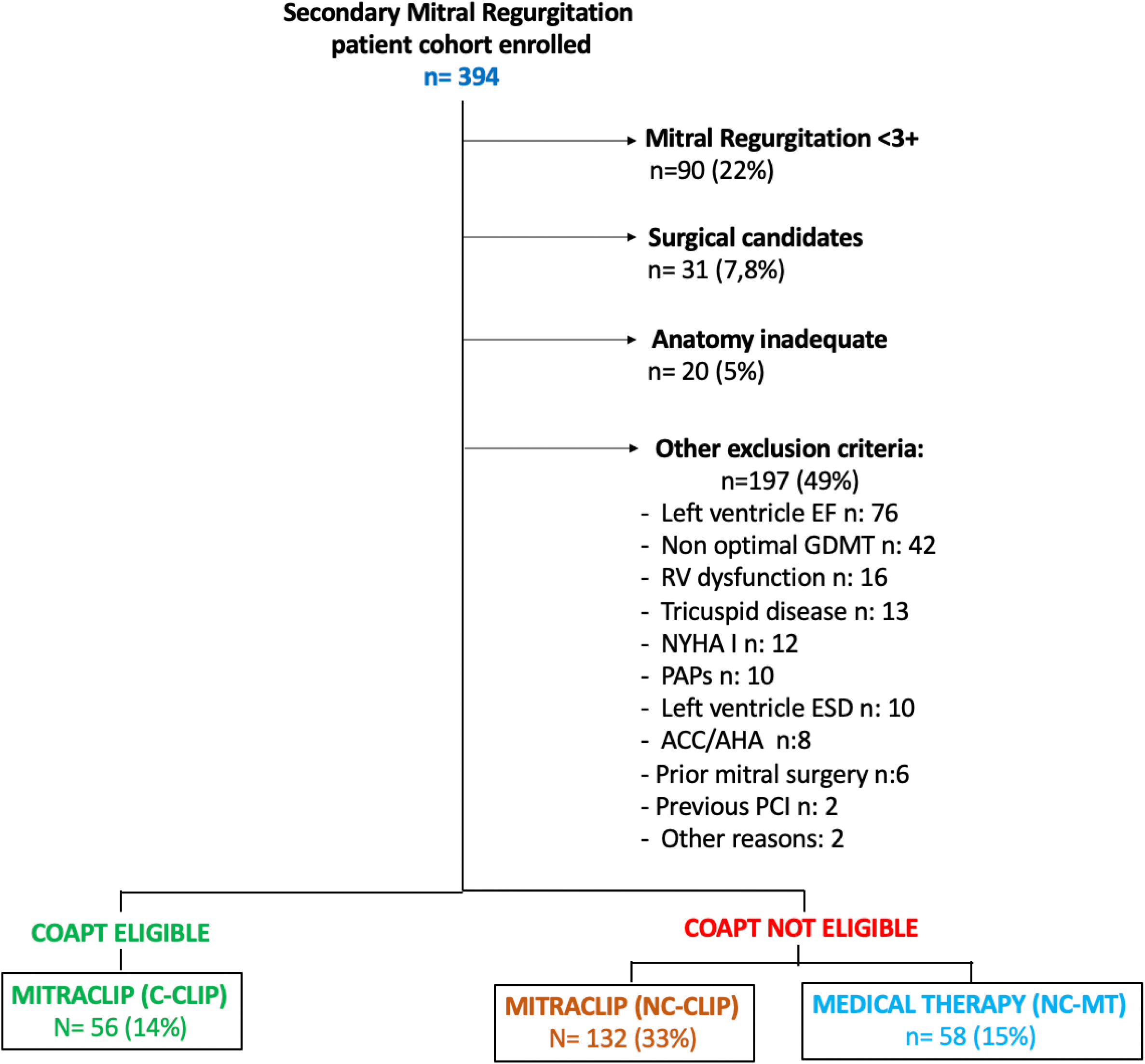
COAPT Inclusion and Exclusion criteria flowchart. COAPT Inclusion and Exclusion criteria flowchart. CABG: coronary artery bypass grafting; EF: ejection fraction; LVAD: left ventricle assist device; LVESD: left ventricle end systolic diameter; MR: mitral regurgitation; NYHA: New York Heart Association; sPAP: systolic pulmonary artery pressure; RV: right ventricle; TMVR: transcatheter mitral valve replacement.

90 (22%) patients were excluded for evidence of FMR ≤ 2+: among them, 45 (50%) were discharged with adjusted medical therapy while 27 (30%) underwent a non-mitral procedure, 10 (11%) underwent a mitral procedure in association with another intervention and 8 (8,8%) underwent an isolated mitral procedure (1 surgical, 3 MitraClip).

Another 31 patients (7,8%) were found to be good surgical candidates and were treated accordingly.

Further 197 were excluded for the following reasons: 76 (19%) were excluded for LVEF <20% or >50%, 42 patients (10,6%) were excluded for non-optimized GDMT, 20 (5%) for unfavourable mitral valve anatomy, 16 (4%) were not considered eligible as they presented moderate/severe RV dysfunction. Moreover, 12 (3%) were excluded since presented NYHA functional class I, 13 patients (3.2%) showed a tricuspid regurgitation (TR) requiring intervention, 10 patients (2.5%) were excluded because of sPAP >70 mmHg and 10 (2.5%) because they presented a left ventricle end-systolic diameter (LVESD) >70 mm. Moreover, 8 (2%) patients were excluded being HF stage D and 4 (1%) were excluded because of prior mitral valve surgery. Finally, 2 (0.5%) were not considered due to recent PCI treatment and 2 (0.5%) for other reasons.

Among the 338 patients excluded due to the aforementioned criteria, 132 (33%) underwent MitraClip procedure (MC-NC group), 58 (15%) were discharged after optimization of medical therapy (MT-NC group) and 11 patients (2.7%) underwent transcatheter mitral valve implantation (TMVI).

Only 56 patients overall (14%) would have been fully eligible for the COAPT and all of them underwent percutaneous edge-to-edge repair with MitraClip (MC group).

**Table 1** shows the baseline patients characteristics of study groups. Patients with MitraClip outside the COAPT criteria were similar/sicker compared to COAPT-like MitraClip patients. MC-NC patients were also sicker than MT-NC patients: they had larger LV end-diastolic volume index (p=0.001), worse NYHA functional class (p < 0.001), higher prevalence of CKD (p=0.001), lower TAPSE (p=0.001) and higher TR (p=0.02).

**Table 1.**
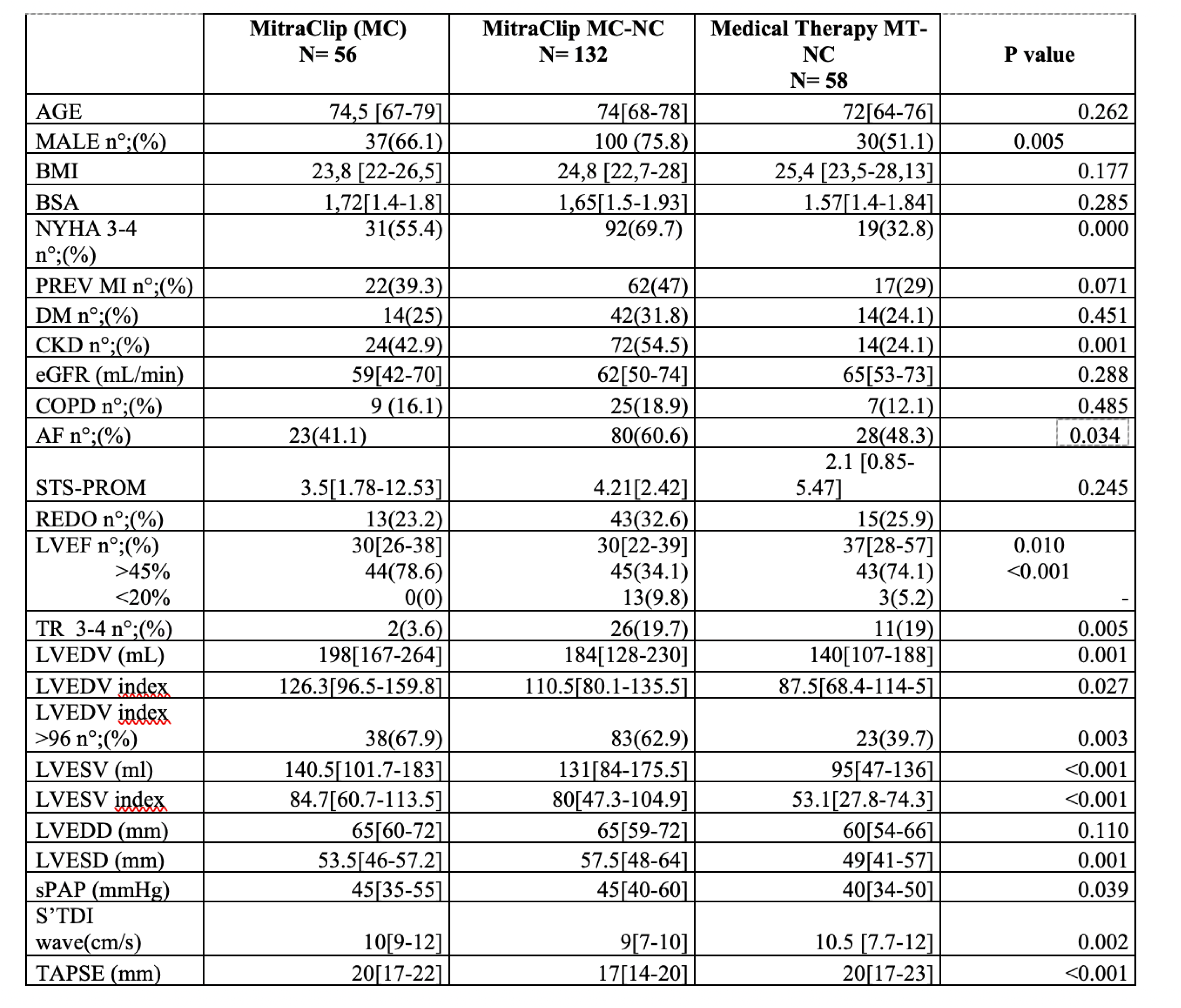
Baseline characteristics of the study population. Values are expressed as mean ± standard deviation; % (n/N) or median [interquartile range]. Bold values indicate statistically significant values. BMI: body mass index; BSA: body surface area; DM: diabetes mellitus; COPD: chronic obstructive pulmonary disease; CKD: chronic kidney disease; eGFR: estimated glomerular filtration rate; LVEF: left ventricular ejection fraction; LVEDD: left ventricle end diastolic diameter; LVEDV: left ventricle end diastolic volume; LVESD: left ventricle end systolic diameter; LVESV: left ventricle end systolic volume; MI: myocardial infarction; NYHA: New York Heart Association sPAP: systolic pulmonary artery pressure; S’ TDI: systolic wave of tissue Doppler imaging; STS: society of thoracic surgeons; TAPSE: tricuspid annular plane systolic excursion; TDI: tissue doppler imaging; TR: tricuspid regurgitation.

### Follow-up outcomes

Follow-up was 99.8% complete, with median follow-up time of 4 years.

#### Survival

Figure 1 reports raw 4-year survival percentages among different study groups according to treatment received.

**Figure 1.**
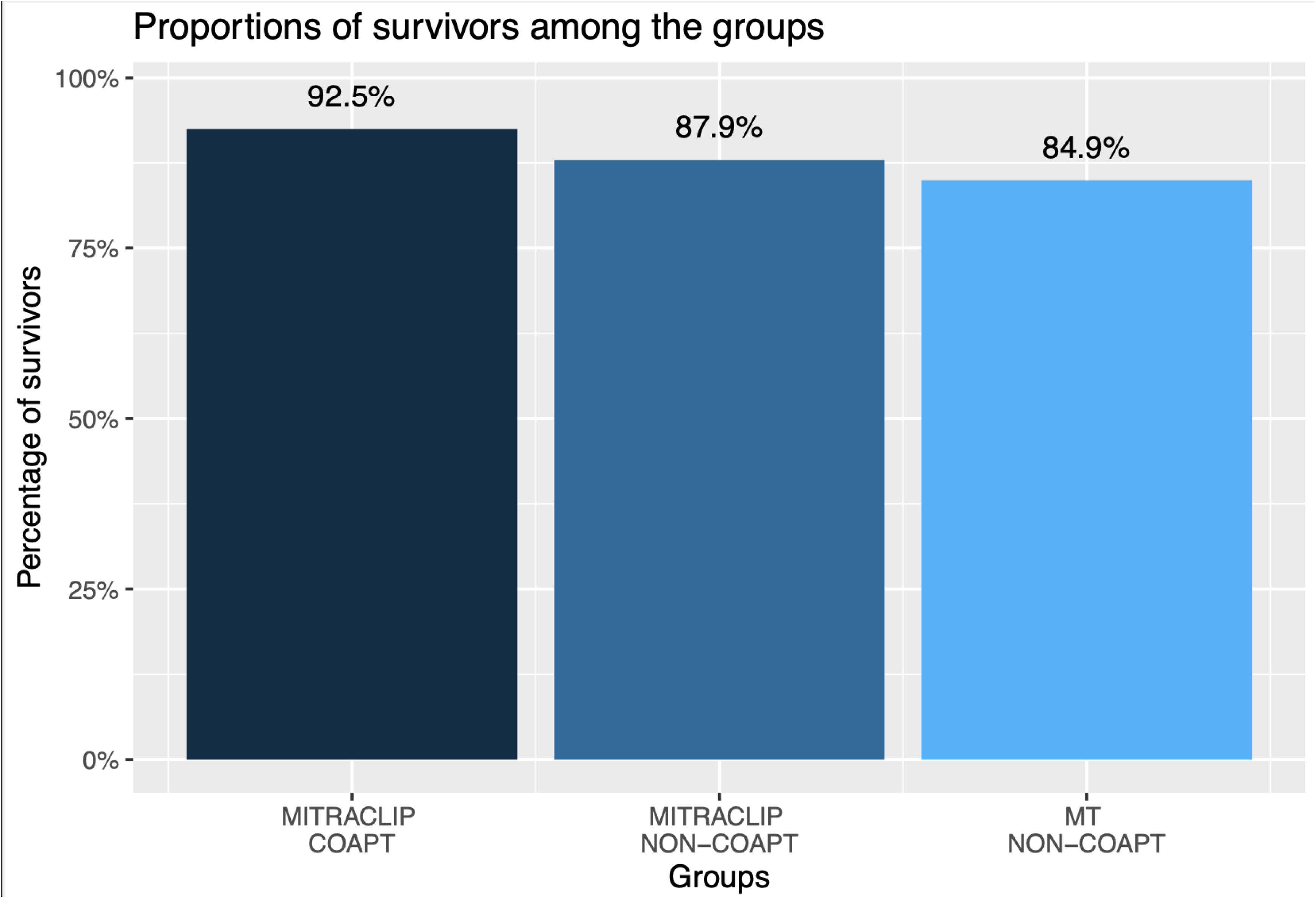
4-year survival percentages among different study groups according to treatment received. Plot of the 4-year overall survival among different study groups according to the treatment received.

The UN-Weighted 4-year survival was similar between MC and MC-NC patients (p=0.57) Figure 2.

**Figure 2.**
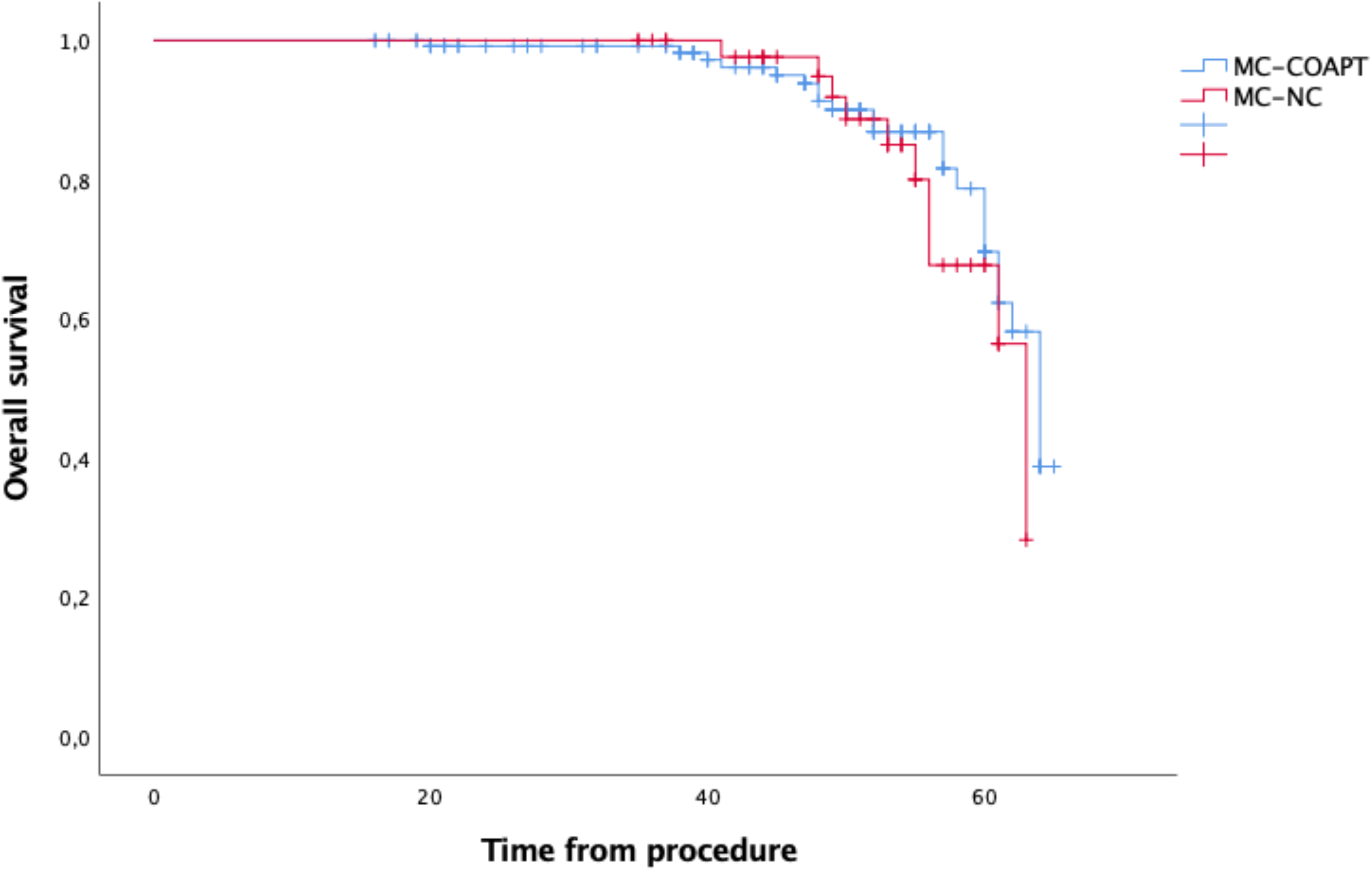
Unweighted Kaplan-Meier curve for 4-year all-cause death. Plot of the occurrence of all-cause death between MitraClip COAPT-eligible (M-C) and MitraClip COAPT-ineligible (M-NC) and (p=0.57)

Among NON-COAPT patients, weighted 4-year survival was higher in patients who received MitraClip compared to those who were left in optimized medical therapy (91.5% (CI: [0.864, 0.96] vs 71.8 % (CI = [0.509, 0.926]) respectively, p=0.027), Figure 3.

**Figure 3.**
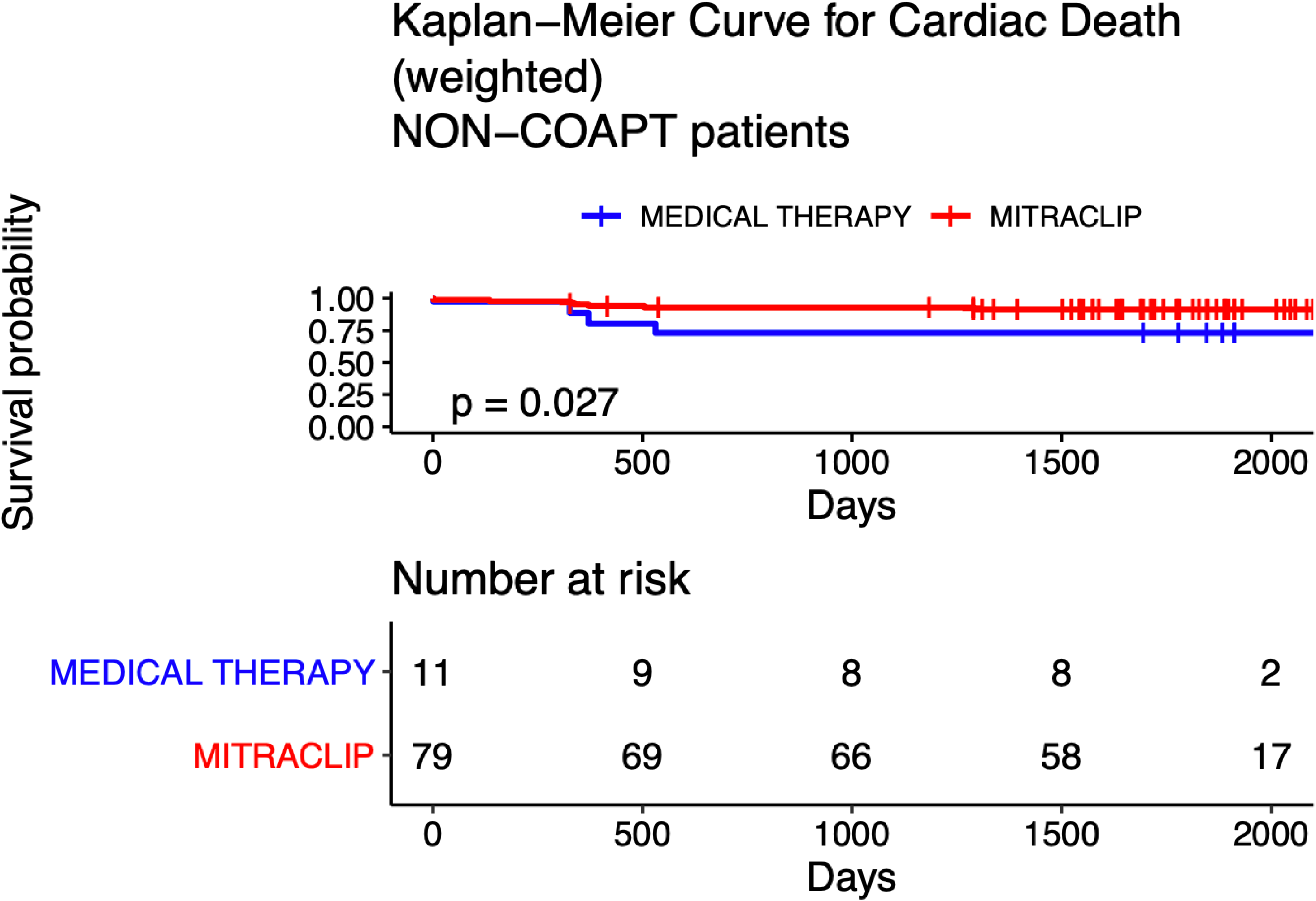
Weighted Kaplan-Meier Curve for 4-year all-cause death. Survival plot of the occurrence of all-cause death between COAPT-ineligible patients treated with MitraClip or medical therapy, with significant difference between the groups (p=0.027).

Predictors of death at 4 years in COAPT-ineligible patients at univariate analysis were age, being a smoker, having beta-blockers in therapy, previous HF hospitalization and low pre-op LVEF, **Table 2**. No variables were significant in multivariate analysis.

**Table 2.**
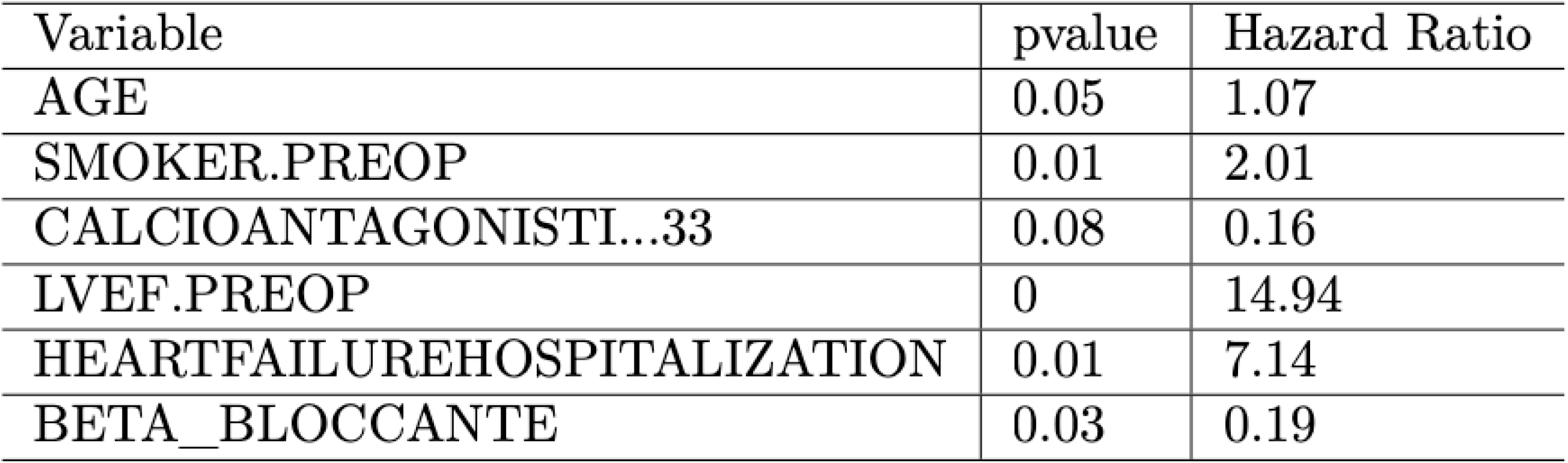
Predictors for 4-year cardiac death.

#### MR recurrence

4-year recurrent MR>=3+ in MitraClip patients was 17,2% (CI: [0.101, 0.238]), while in Medical therapy group was 40% (CI = [0.248, 0.522]), p 0.0019; Figure 4.

**Figure 4.**
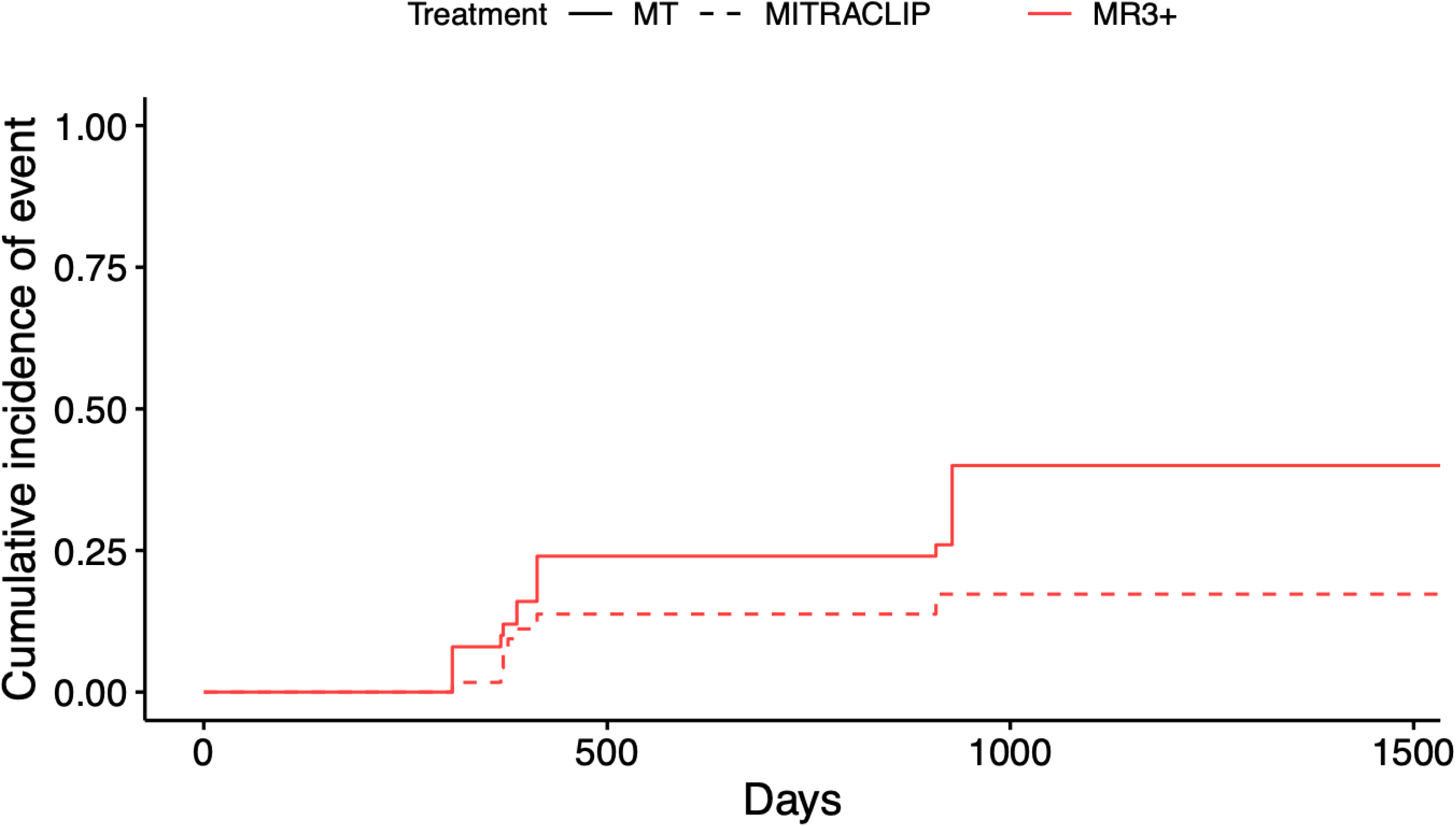
Cumulative incidence function of MR>=3+ recurrence at 4 years. Plot of the occurrence of residual/recurrent MR>=3+ at 4 years between COAPT ineligible patients treated with MitraClip and medical therapy group, with significant difference between the two groups (p=0.019).

Similar results were obtained if considered 4-year recurrent MR>=2+, p=0.0067; Figure 5.

**Figure 5.**
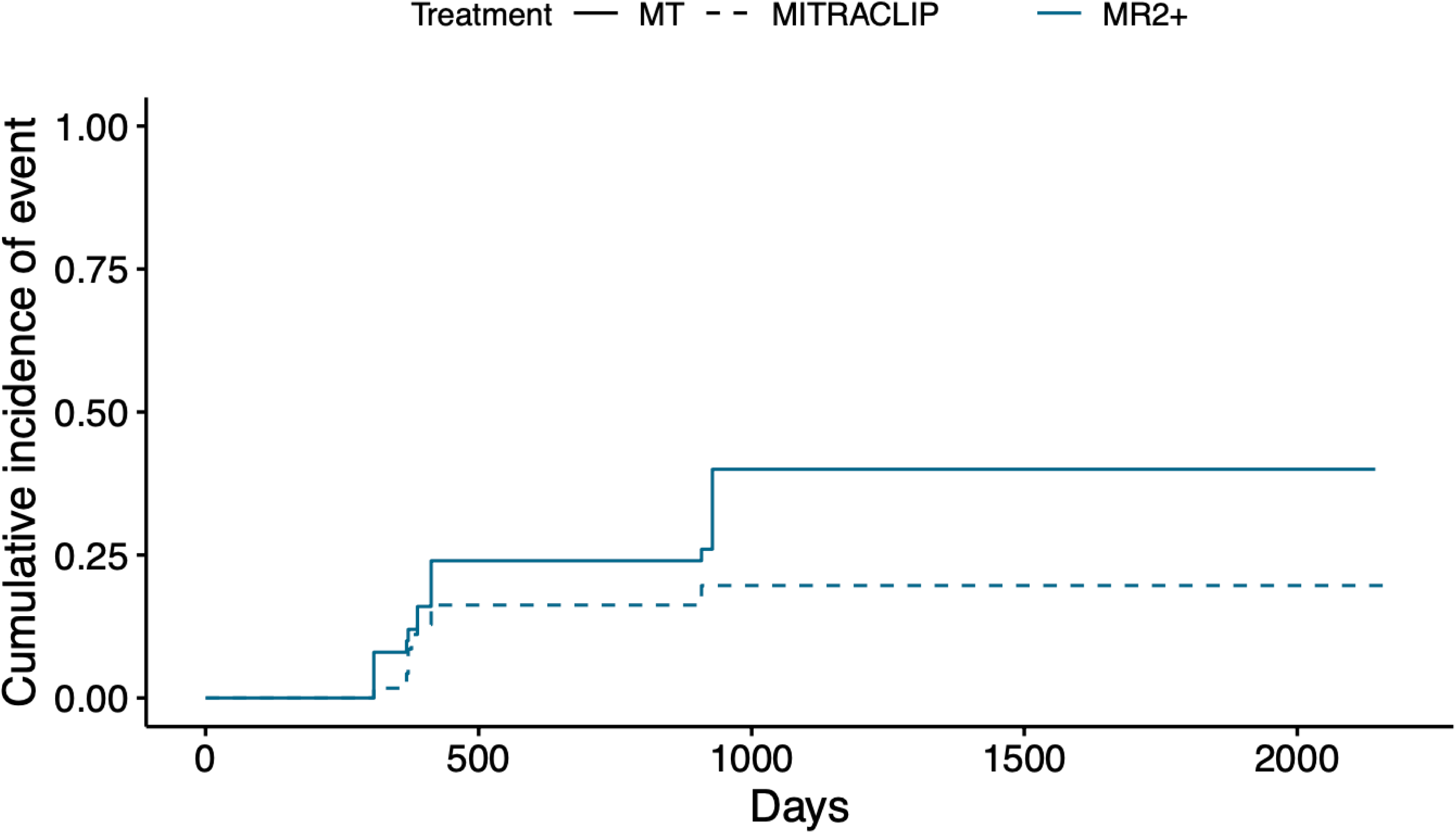
Cumulative incidence function of MR>=2+ recurrence at 4 years. Plot of the occurrence of residual/recurrent MR>=2+ at 4 years between COAPT ineligible patients treated with MitraClip and medical therapy group, with significant difference between the two groups (p=0.0067).

#### NYHA class status

NYHA at follow-up for MitraClip patients was lower than the one for patients following medical therapy, with again a p-value < 0.0001; Figure 6.

**Figure 6.**
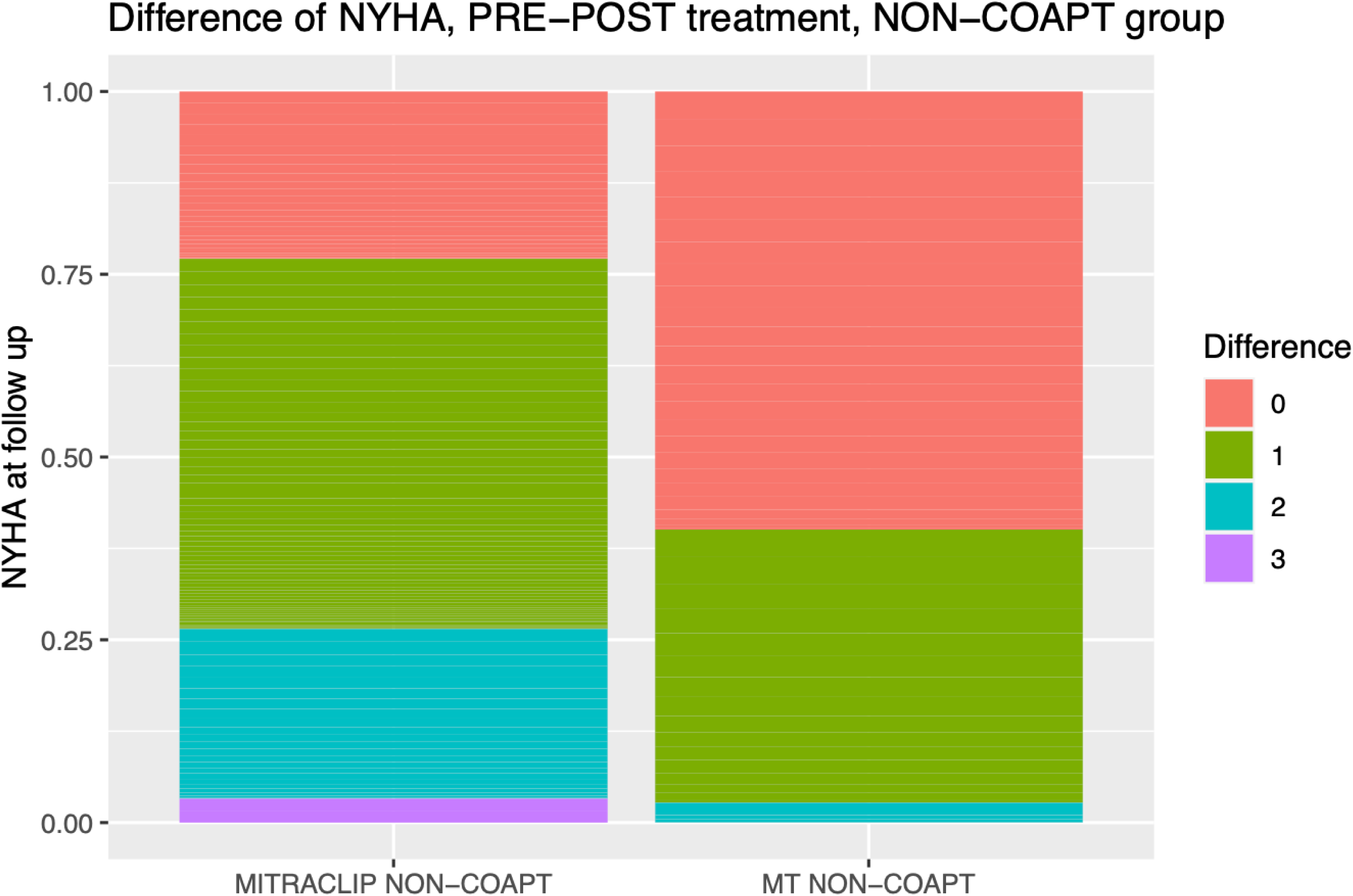
Wilcoxon non-parametric test to examine the NYHA class change. A Wilcoxon non-parametric test for samples to test the change in the mean of NYHA preop and at follow-up in the NON-COAPT group.

## Discussion

The main findings of the present study are that in a wide patient population referred for treatment of FMR in a real-world setting:

1. Only a minority of patients presented within the COAPT selection criteria;
2. In patients outside COAPT criteria but who were confirmed to have MR>=3+ and were not surgical candidates, after 4-year MitraClip provided similar survival to that observed in COAPT eligible patients and improved survival compared to medical therapy.

The debate about COAPT vs MITRA-FR trials over the last years usually revolves around how in the COAPT the hearts were more strictly selected to be “better” and the MR to be more “critical” compared to those in the MITRA-FR(5), (7), (8,9). Such concepts, in turn, have been recently brought into question by a post-hoc analysis of MITRA-FR data, which despite focusing on LV remodeling and MR quantification parameters, could not identify any subset of patients that might have benefited from transcatheter repair with Mitraclip(10).

The present study aimed to provide real-life insights in this clinical scenario and confirmed that COAPT selection is not easily reproducible in daily practice, with only 56 patients (14%) eligible out of the entire FMR patient population referred to our centre over 4 years. While such number seems low, it actually becomes surprisingly high when compared to the COAPT three top enrolling centres who recruited respectively 46, 30 and 29 patients over 4.75 years (approximately 7-8 patients per year). Such discrepancy may be explained by several reasons, including the time needed for patient reassessment after optimization of medical therapy, the RCT two screening committees time schedule, the will to distribute RCT enrolled patients across multiple sites (limiting peaks at more experienced ones). On the other hand, the presence of some blurred and unspecified selection criteria such as the mitral anatomy which could have caused some patient selection to remain submerged.

Compared to the RCT device arm, our real-life COAPT population showed similar baseline features, although worse LV dilatation must be noted (LVEDVi 101 mL/mq vs 124 mL/mq, respectively). Despite the meticulous application of all officially reported COAPT criteria, we still cannot know if our selected patients were true peers of RCT accepted cases. Mitral anatomy selection could have played a role in this regard. Indeed the above-observed discrepancy in LV volumes may subtend a worse mitral anatomy in our real-world population compared to the RCT. Unfortunately, no official data concerning the specifics of mitral anatomy from the COAPT trial are available. The judgment of anatomical feasibility is clearly highly dependent of operators experience and confidence and can easily explain significant differences in patients selection across centres and studies. Indeed, we still lack a standardized unified method to describe anatomy suitability for MitraClip; using the German proposal by Boekstegers et al. (11) in our treated population among all 188 MitraClip procedures only 34 patients (18%) presented an *optimal* valve morphology, while most were judged as *conditionally suitable* (n=152; 80,8%) or *unsuitable* (n=2; ≍1,6%). Even among our 56 COAPT eligible cases, 11 (19%) patients presented *optimal* valve morphology, while 45 (80%) were considered *conditionally suitable, due to frequent Carpentier IIIb MR mechanism*. The unspecified anatomy criteria in COAPT trial did not allow precise retrospective selection of our cohort. In turn this could also partially explain why our COAPT and non-COAPT patients look closer than expected.

Few other considerations are worth to be noted:

- 1/5 of patients referred to receive treatment in our practice did not show “critical” MR. This is a common finding with secondary MR and underlines its dynamic nature(12). In dubious cases, stress echocardiography may be a valuable tool to rule out borderline situations(6). Notably, the survival of these patients with MR<=2+ was actually disappointing and the explanation of this may be actually two-fold: on one hand, fluctuant MR can be disguised, on the other, secondary MR remains by definition not the primary heart disease.
- in the present experience, no significant difference in follow-up survival was observed between COAPT eligible and non-COAPT patients treated with MitraClip and as a matter of fact the two groups were surprisingly similar also in terms of baseline features, although a trend towards worse profile in non-COAPT patients could be observed in several variables. The small numbers, especially in the COAPT group, likely impair most statistical analysis between these two groups and therefore no reliable conclusion can be drawn in this regard. In a recently reported real-life study, COAPT-like patients treated with MitraClip had a significantly lower mortality and less HF hospitalizations compared to non-COAPT patients(13). A higher proportion of patients (n=61; 51%) were eligible for COAPT in this study compared to our experience, but all these patients had already been preselected by Heart Team to be treated with MitraClip after being on GDMT for 3 months, resulting in an overestimation of COAPT eligibility rate in the true all-comer population.
- Real-world patients treated with MitraClip even outside COAPT criteria showed better 4-year survival compared to non-COAPT patients left in medical therapy, despite a tendency towards worse baseline profile in MitraClip patients remained even after weighting. This finding raises some questions about whether it is truly convenient to delay FMR correction to later stages of medial therapy and may support the use of MitraClip even for patients not completely fulfilling COAPT criteria. Larger numbers and longer follow-up will be needed however to validate this finding. The opportunity intervene on patients outside the COAPT and who still have room for drug optimization must be carefully weighted and tailored by the Heart-Team to each and every single case. On the other hand it must be underlined that late timing of intervention is already a recognized mistake in surgery, associated with worse outcomes(14). Earlier timing of FMR correction may be beneficial to selected patients, similarly to other valvular heart disease (15),(16). Therefore, a major goal of Heart Team is to identify which burden of FMR is going to be critical in different stages of HF. An observational study addressing the prognostic impact of FMR in a large contemporary HF population identified a subset of intermediate phenotype patients, characterized by a moderate impairment of functional status and LV dysfunction, where FMR remains an independent predictor of outcome despite optimized medical therapy and which could likely benefit from transcatheter repair to improve survival(17).

## Limitations

The present was a retrospective single-centre study including a small cohort population and a short follow-up, therefore its findings should be considered hypothesis generating and will require further data to be confirmed. While assessment of medical therapy was carefully performed, retrospective evaluation remains a significant limitation. Lack of EROA values for all patients included in the study did not allow greater characterization of FMR and a more precise comparison with COAPT trial population. Also, some of the original COAPT exclusion criteria are not fully specified and therefore their interpretation may have led to different patient selection compared to the original trial: e.g. RV dysfunction and mitral anatomical selection. Finally, despite propensity matching, there may be residual confounding factors influencing overall survival between groups that could not be completely accounted for.

## Conclusion

In the present retrospective, single-centre, real-world study only a minority of patients referred to treatment of FMR presented within the COAPT trial selection criteria. The main reasons for ineligibility included absence of MR>=3+, eligibility to surgery, non-optimized GDMT, and inadequate LVEF. Despite a worse baseline clinical profile, patients outside COAPT criteria treated with MitraClip experienced improved 4-year survival compared to those left in medical therapy. While these findings require further validation on longer follow-up, the numerous patients currently referred to treatment outside the COAPT criteria should not be denied intervention but should receive a tailored Heart-Team evaluation. Further refinement of patients selection to transcatheter mitral valve interventions remain needed.

## Clinical perspectives

### Competency in medical knowledge

The COAPT was the first ever trial demonstrating a survival in benefit in patients with FMR treated with MitraClip plus GDMT compared to GDMT only. The results of the present study suggest that only a minority of patients presenting with FMR in a real-life scenario could be COAPT-eligible but that patients presenting outside COAPT criteria can still benefit from MitraClip, with an improved survival compared to medical therapy and a similar survival compared to COAPT-like patients.

### Translational outlook

While these findings require further validation, the numerous patients currently referred to percutaneous repair outside the COAPT criteria should not be denied intervention and should receive a tailored Heart-Team evaluation. Further refinement of patients selection to transcatheter mitral valve repair and of real-world applicability of COAPT criteria and results are needed.

## Conflict of Interest Disclosures

Francesco Maisano: Grant and/or Research Support Abbott; Medtronic; Edwards Lifesciences;

Biotronik; Boston Scientific Corporation

Consulting fees, Honoraria Abbott; Medtronic; Edwards Lifesciences; Swissvortex

Perifect; Xeltis; Transseptal solutions; Cardiovalve

Royalty Income/IP Rights Edwards Lifesciences (FMR surgical annuloplasty);

4Tech (tricuspid catheter interventions)

The other authors do not have COI.

## Data Availability

The data collected are available on demand

## Abbreviations

FMR: functional mitral regurgitation
GDMT: guidelines directed medical therapy
HF: heart failure
LVEF: left ventricle ejection fraction
LVEDV: left ventricle end diastolic volume
LVESV: left ventricle end systolic volume
MC: MitraClip in COAPT patients
MNC: MitraClip in Non COAPT patients
MTNC: Medical therapy in Non COAPT patients
TAPSE: tricuspid anular plane systolic expansion

## Acknowledgments

none

## Funding, grant/award info

None

Data sharing are available upon request

The San Raffaele Hospital Institutional Ethic Committee approved this study on mitral regurgitation and tricuspid insufficiency 125/INT/2022 and waived individual consent for this retrospective analysis. And the study must comply with the Declaration of Helsinki. The research protocol must have been approved by the locally appointed ethics committee and informed consent must have been obtained from subjects (or their guardians).

